# How are social and economic needs assessed and addressed in mental health services? A service evaluation of the largest mental health trust in the UK

**DOI:** 10.1101/2025.05.12.25327421

**Authors:** Anna Greenburgh, Helen Baldwin, Hannah Weir, Zara Asif, Dionne Laporte, Mark Bertram, Achille Crawford, Gabrielle Duberry, Shoshana Lauter, Brynmor Lloyd-Evans, Cassandra Lovelock, Jayati Das-Munshi, Craig Morgan

## Abstract

There is an urgent need for services to address the social and economic adversities which contribute to the aetiology and outcomes of mental health problems. However, the implementation of interventions to do so is inconsistent, and entrenched cycles of poor mental health and social exclusion persist. We conducted a service evaluation survey of 28 staff working across community and inpatient teams, enhanced by a series of in-depth case studies, in the largest NHS mental health trust in the UK to explore how social and economic needs are currently assessed and addressed. We found that assessment for social and economic needs varied across different domains; for example, family relationships were more consistently assessed than domains such as education and income. A range of support is available and provided by a patchwork of teams, including Community Mental Health Teams, other NHS teams, Local Authority staff, and many third sector organisations. However, what support is available is severely restricted and respondents highlighted a lack of adequate support in every domain we considered – employment, education and training, social participation and connectedness, family relationships, community support, social security, debt, income, housing, and trauma and victimisation, as well as additional domains including healthy eating, sex and relationships, and activities of daily living. Nevertheless, our case studies illustrate examples of approaches to addressing social and economic needs to improve outcomes for people with mental health problems.

## 1. Introduction

People experiencing mental health problems often are caught in entrenched cycles of social disadvantage, worsening mental health, and poor experiences with services. People facing socioeconomic adversities are less likely to access timely primary and secondary care, more likely to access crisis services, and less likely to experience improved outcomes once care is received (Barnett et al., 2023). These inequalities are directly relevant to the ethnic inequalities observed in health care systems such as those in the UK and USA. That is, as people with mental health problems from Black and other minoritised ethnic communities are more likely to experience social and economic disadvantage (Chilman et al., 2024; Morgan et al., 2008, 2017), and this disadvantage restricts effectiveness of treatment. Services that do not address these adversities risk sustaining systemic inequalities.

Accordingly, it is increasingly understood that services must assess and address the social and economic needs faced by people with mental health problems. Tools to assess and track social inclusion outcomes and improve person-centred care have been developed, such as the SINQUE (Eager et al., 2024) and DIALOG+ (Priebe et al., 2015). Equally, interventions to improve social and economic circumstances have been evidenced, with particular support being found for supported housing and supported employment interventions, namely Housing First and Individual Placement and Support (Barnett et al., 2022; Greenburgh et al., 2025; Killaspy et al., 2022). However, progress is still needed in this field, as services tend to focus on medication and risk management rather than addressing social and economic needs. Indeed, people in contact with mental health services report lack of support with fundamental needs, leading to loss of trust in services (Lambri et al., 2012), and there is a notable lack of evidence for interventions to ameliorate many social and economic problems, such as debt and social security, which worsen mental health (Baldwin et al., 2025; Barnett et al., 2022; Killaspy et al., 2022).

In the UK, implementation of methods to assess and address social and economic challenges of people with mental health problems are underdeveloped, inconsistent and opaque. NHS mental health trusts spend more on inpatient care than community-based care such that the majority of budgets are spent on a minority of people in need (Gilburt & Mallorie, 2024), where these individuals experience have a very high level of need; however, higher community spend is associated with reduced acute demand across the NHS (Gorham & Wood, 2023).

NHS staff responsibilities regarding social inclusion are unclear. Indeed, it is local councils, rather than the NHS, which have legal responsibilities with regard to prevention and wellbeing promotion, as outlined by the Care Act of 2014; and the Voluntary, Community and Social Enterprise (VCSE) sector receives its funding from local authorities - not the NHS - in relation to providing mental health support (Mental Health Social Care Policy & and Oversight Group, 2022). Nevertheless, support with social and economic needs is often provided by a range of NHS and non-NHS staff, including social workers in multi-disciplinary Community Mental Health Teams (CMHTs), other NHS staff such as occupational therapists, local authority staff, and VCSE staff. Integrated Care Systems have been introduced with the aim to improve relationships and partnerships between these teams and sectors and form part of the NHS Long Term Plan (2019), which has also seen the initiation of the Community Mental Health Framework for Adults and Older Adults. However, the extent to which this framework is being implemented, and consequently its effectiveness on reducing social exclusion of people with mental health problems, remains unclear.

## 2. Aims

We conducted a service evaluation survey of staff working in mental health teams in South London and Maudsley NHS Foundation Trust to explore how social and economic needs are assessed and addressed (see SI for aims in full).

## 3. Methods

### 3.1 Service context

South London and Maudsley NHS Foundation Trust (SLaM) is the UK’s largest mental health trust, serving a population of about 1.3 million people. The catchment area consists of four boroughs, Southwark, Lambeth, Croydon and Lewisham, which are among the most income-deprived in the UK (16.2%,15.3%, 13.6%,16.4%, income-deprived, respectively (ONS, 2021)). Users of SLaM services specifically face high degrees of social and economic need: an estimated 83% of people in contact with SLaM services between 2005-2020 were in receipt of benefits at some point in this period (Stevelink et al., 2023); 84.9% of people with severe mental illness in contact with secondary mental health services in SLaM have had periods of unemployment (based on records extracted in 2020 (Chilman et al., 2024)). Many people in contact with SLaM services also experience loneliness – and this predicts outcomes such as subsequent crisis episodes and emergency presentation (Parmar et al., 2024). There is also considerable disability and poor health which cooccur with economic inactivity and are associated with longer illness duration (Cybulski et al., 2024). Recent initiatives to transform services have been launched, such as the 24/7 community mental health service in Lewisham. Nevertheless, little is known about how social and economic needs are currently assessed and addressed more broadly across SLaM as a whole.

### 3.2 Procedure

We conducted a service evaluation survey recruiting staff working in mental health services across SLaM. We worked with members of our Advisory Board of people with lived experience of mental health problems and services, service providers, clinicians, academics and commissioners to iteratively codevelop this survey. The service evaluation was registered and approved via MAUD (ID 403). We recruited a convenience sample by circulating the survey through official SLaM communication channels, presentations to teams in SLaM, and dissemination across our networks and the networks of our Advisory Board members. Data collection took place from February to September 2024. Informed consent was obtained from all respondents, participation was anonymous, and no personal data was collected at any stage. The survey was completed using Qualtrics whereby respondents took part through an anonymised link.

In tandem, we produced case studies of NHS services and related third sector organisations working in the SLaM catchment area to improve social and economic outcomes of people with mental health problems. These were written through conducting site visits (minimum 1 per case study), interviews and meetings staff members of these teams. We then produced a narrative summary of the work of each group, describing the needs they addressed of people in contact with their services, the positive impacts they had achieved, and the challenges they faced.

### 3.3 Measures

The co-developed service evaluation survey comprised 41 questions and took approximately 10-15 minutes to complete (see SI for more details).

### 3.4 Data analysis

Data were summarised using frequency counts and simple descriptive statistics using R studio ((*RStudio: Integrated Development for R*, 2020) and data visualisations were created using ggplot2 (Wickham, 2016), likert (Bryer & Speerschneider, 2016), and hrbrthemes (Rudis et al., 2024) packages. Qualitative data (e.g. noting types of support available; issues raised concerning support) were listed verbatim or narratively summarised.

## 4. Results

### 4.1 Participants

Twenty-eight participants working in adult services took part in the survey (see SI for roles). Of these, 21 respondents reported that they worked in a community-based team (e.g. CMHT; EIP Community Team; Early Onset Community Team) and 7 worked in other types of teams (e.g. Acute hospital liaison psychiatry) (see SI for further detail on team and caseloads).

### 4.2 Aim 1: to explore the extent to which social and economic needs of people in contact with mental health services are routinely assessed by teams

**Table 1.** The types of support available in each domain, as listed by survey respondents.

| Community support | Debt | Education and training |
| --- | --- | --- |
| ◇ Support groups<br>◇ Football and walking groups<br>◇ Gardening projects<br>◇ Art and music therapy<br>◇ Crisis support<br>◇ Clubhouses<br>◇ Community centres<br>◇ Activities within supported housing or residential/nursing placements<br>◇ Peer support | ◇ Safeguarding<br>◇ Applications to court of protection<br>◇ Assessment and support to manage finances<br>◇ Occupational therapy<br>◇ Specialist advice<br>◇ Debt relief from reputable organisations<br>◇ Debt relief programmes<br>◇ Referral to debt agencies<br>◇ Provision of information from Government websites<br>◇ Support with making appointments at Citizens Advice Bureau<br>◇ Support with telephone calls and letters if required to individual organisations | ◇ Recovery College<br>◇ Referral to skills development services<br>◇ Local colleges<br>◇ Online courses<br>◇ Training schemes<br>◇ Work placements<br>◇ Adult learning and training services<br>◇ Education courses<br>◇ Peer Support<br>◇ One-to-one support from Vocational Workers<br>◇ IPS |

| Employment | Family relationships | Housing | Income |
| --- | --- | --- | --- |
| ◇ Individual Placement and Support (IPS)<br>◇ Referral skills development services<br>◇ Local workplace opportunities<br>◇ One-to-one employment support<br>◇ Careers advice<br>◇ Specialist vocational support<br>◇ Support with occupational health queries<br>◇ Signposting to relevant services<br>◇ Paid work training programme | ◇ Network meeting clinic<br>◇ Family therapy<br>◇ Family interventions in psychosis<br>◇ Informal and formal carer/family support<br>◇ Carers assessments<br>◇ Safeguarding, early help<br>◇ Care in line with the Triangle of Care model | ◇ Liaising with Local Authorities regarding housing issues<br>◇ Tenancy management support<br>◇ Supporting letters<br>◇ Inhouse advice for homelessness<br>◇ Information from local charities<br>◇ Support finding placements<br>◇ Housing referrals<br>◇ Homeless Person's referrals<br>◇ Funding panels, individual referrals to specialist placement<br>◇ Signposting to housing services<br>◇ Support with application to mainstream housing and supported housing options<br>◇ Street homeless applications | ◇ Safeguarding, applications to court of protection<br>◇ Assessment and support to manage own finances<br>◇ Support accessing benefits and obtaining banks<br>◇ Advice from external organisations<br>◇ Referral to local authorities<br>◇ Referral to financial advisor<br>◇ Support with making telephone calls and completing forms |

| Social participation and connectedness | Social security | Trauma and victimisation | Other |
| --- | --- | --- | --- |
| ◇ Support groups<br>◇ Support and skills building including with relationships and friendships<br>◇ Local community groups, cinema trips, walking groups, volunteering, social groups<br>◇ Referral to skills development services<br>◇ Signposting to community organisations<br>◇ Referral to NHS-led groups for activities such as gardening<br>◇ Wellbeing hubs<br>◇ Coffee with Support Workers<br>◇ Peer Support group visits to parks, art galleries, museums, theatres and restaurants | ◇ Benefits advisors<br>◇ Specialist advice<br>◇ Signposting<br>◇ Carer support groups<br>◇ One-to-one benefits support e.g. with appeals, reviews, and applications | ◇ Psychological therapy,<br>◇ Group interventions<br>◇ Specialist trauma support<br>◇ Safeguarding<br>◇ Vulnerable adults' assessments<br>◇ Risk assessments<br>◇ Advice<br>◇ Referrals to specialist organisation<br>◇ Access to community groups<br>◇ Trauma-Focused CBT<br>◇ Trauma-informed services<br>◇ Domestic abuse support<br>◇ Peer support<br>◇ Carer's groups | ◇ Culturally specific social support<br>◇ Spiritual support<br>◇ Religious groups<br>◇ Support with sex and relationships<br>◇ Parenting support<br>◇ Carers groups<br>◇ Social prescribing<br>◇ Peer groups<br>◇ Supporting social networks and general functioning<br>◇ Substance use and addiction support services<br>◇ Service user networks<br>◇ Meditation activities<br>◇ Practical interventions (including supporting someone collect their medications; cooking together, walking together) |

For each domain, at least 60% of respondents (n=15) said that needs were ‘always’ or ‘often’ assessed, and the median number of respondents reporting that needs were ‘always’ or ‘often’ assessed for each domain was 21. Assessment consistency was heterogeneous across domains (see Figure 1).

**Figure 1.**
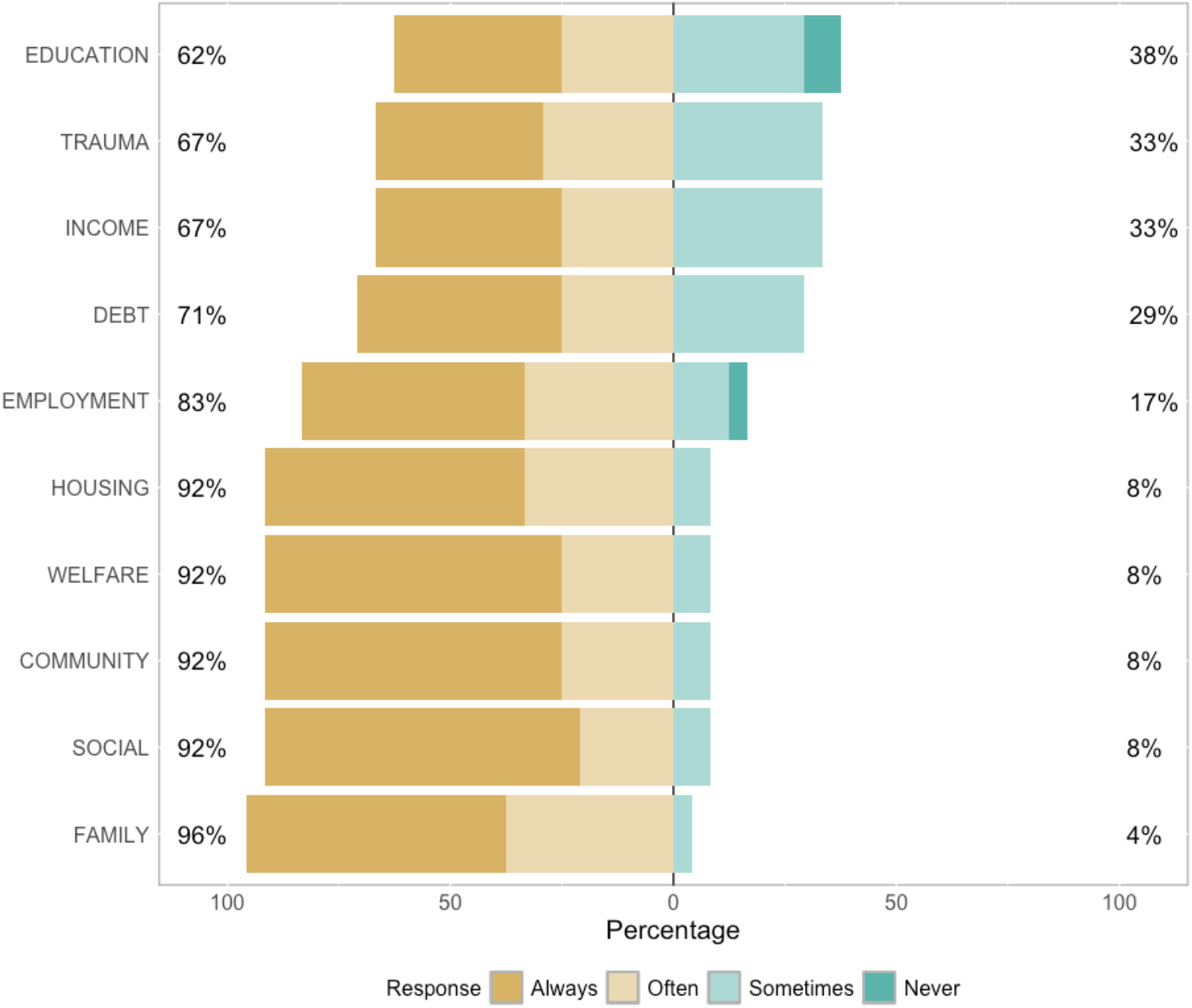
Reported extent to which service-users’ needs across 10 core domains are assessed within respondents’ teams

### 4.3 Aim 2: to better understand what services and support are provided in relation to the social and economic needs of people in contact with mental health services

#### Examples of excellent practice

The below anonymised case studies of examples within SLaM NHS and related third sector organisations of interventions, describe four services and groups working to improve social and economic outcomes for people with mental health problems (See SI for full narrative summaries of challenges and successes for each service).

The social security advice service offers expert welfare benefits advice, casework, and advocacy. The team works to improve the financial situation and housing security of service users, thereby tackling some of the most important social determinants of mental health problems. Their specialist advice helps service users to navigate often opaque and complex welfare systems facilitating financial safety and security. Indeed, the social security system has become more conditional in recent times and therefore harder to navigate for those experiencing mental health problems, results in worse mental health (Baekgaard et al., 2024; Dwyer et al., 2020). The team works to ensure that service users receive the social security they are entitled to, and they are often able to bring service users out of poverty by doing so.

The culturally appropriate peer support and advocacy service works across the community and in-patient services, to help service users and their families to feel empowered in their recovery journeys. The team finds new, innovative ways of working to address the failures of current mainstream services when it comes to providing care for Black communities and to tackle racial inequalities in mental health services. The service works to empower their clients to both self-advocate and seek support in their recovery journey. The peer advocates work to challenge diagnosis, medication, restrictive practices, and treatments that disproportionately impact Black service users, such as Community Treatment Orders and seclusion. They also offer one-to-one support in both the community and in-service, which includes liaising with different stakeholders in care, family members, friends and clinicians.

The vocational service supports social inclusion for people with mental health problems across the domains of education, employment, training, and volunteering. They provide person-centred support to enable individuals to find their own way forward and engage in vocational activities that are important to them. The service also helps their clients navigate the welfare benefits and housing systems, as the majority face problems in these domains, which need to be addressed before progress can be made with supporting vocational activities. They run many projects including user-led services. The service is open to working-age adults and is accessible through self-referral or referral by a health professional.

The specialised psychological therapies team for people with psychosis comprises a Peer Support Service and a Family and Friends Support Service. The Peer Support Service offers one-to-one support for any person with lived experience who attends the clinic. Peer supporters help clients to become familiar with the process of therapy while they are on the waiting list, and support them to access and engage with therapy, through sharing their own experience and offering practical assistance, for example by accompanying clients to appointments at the clinic when in-person attendance is an obstacle to accessing the help they need. In addition to this one-to-one support, the peer support team run a weekly group meet-up, alternating between a social group and an arts group. This peer support is available to clients after they have been discharged from therapy. The Family and Friends Support Service provides 6 weekly sessions to loved ones of clients attending the clinic. The support provided draws on principles established from family intervention research, and employs materials co-produced with carers. The sessions offer the space for carers to share their experiences, learn more about psychosis and bipolar disorder, receive support with relapse prevention and crisis planning, while also working towards individualised goals to improve their own wellbeing. Staff also assess trauma and presence of PTSD routinely using standardised measures, and signpost attendees to sources of additional support where needed.

### 4.4 Aim 3: to better understand the issues and gaps in provision of support with social and economic needs

Respondents highlighted a lack of adequate support available to people with mental health problems in every social and economic domain. Specific issues raised for each domain were as follows:

#### Employment

The ability for care coordinators to support individuals depends on previous experiences and links with other agencies they’re aware of; the fact that some teams lack access to IPS; and finally, the broad lack of employment opportunities.

#### Education and training

People with mental health problems often have difficulty in accessing a Recovery College as they are oversubscribed and that there is very little in the way of learning opportunities and resources.

#### Social participation and connectedness

Needs around healthy and supportive friendships are not being met. There is a lack of support workers in teams to help individuals with mental health problems engage with social inclusion; a lack of sports, music and arts groups; and a lack of befriending services. The support that is available is often withdrawn too soon: services are quick to reduce support once someone’s mental health has improved regardless of whether they have recovered social connections and other key aspects of their life.

#### Family relationships

More family support is needed. The complexity faced in liaising with social and safeguarding services when supporting with family relationships was noted.

#### Community support

There is a lack of options for community support for people in residential care and high support placements. Difficulty in accessing support for bereavement and loss was also highlighted. Relevant to all domains, survey respondents emphasised the need for sufficient peer support.

#### Social security

Navigating the benefits system is increasingly complex and there is a lack of available benefits advice or direct access to support in the community. One respondent reported that support is often only offered when someone is in hospital. Further, where teams did provide relevant support, this was noted as requiring a huge amount of staff time. Respondents highlighted that specialist external organisations are very busy and have long waiting lists; and that some external organisations are not trained to support this client group and find it difficult to assist people (e.g. if appointments are missed).

#### Debt

There is a broad lack of support available to assist people with mental health problems in managing with debt.

#### Income

There is a lack of specific support available regarding income.

#### Housing

There is a severe lack of housing support and the absence of enough housing to meet need. Respondents highlighted the lack of advice or support and chaotic referral systems. One respondent noted that existing services which are meant to provide support with housing lacked the expertise to do so for this client group. It was also noted that there are not enough higher and medium high supported accommodations in mainstream housing for adults under 60s.

#### Trauma and victimisation

Accessing support for trauma is difficult, as there are many barriers due to eligibility criteria, and there is a general lack of support due to vacant psychologist and psychology assistant posts.

#### Further needs

Many further areas were listed as lacking sufficient support beyond those listed in the above domains. This included support with Activities of Daily Living, peer support, access to community hubs and day centres, social prescribing, health champions, support and learning opportunities for activities such as cooking and healthy eating, support with affordable and healthy lifestyles, sex and relationships support, long-term therapy for people with anxiety and personality disorder diagnoses, support for autistic people and people with learning disabilities, access to regular appointments with professionals assisting socialising, counselling, and support with self-care.

### 4.5: Aim 4: to identify whether available support is provided directly by community mental health teams or via referral to others

In all domains, service users were reported to be supported by a patchwork of teams and groups. In 5 out of the 10 domains, any one given type of CMHT team member (generic, specialist, care coordinator, other) was reported as the most likely provider of support (employment, social participation and connectedness, family relationships, income (equal to other community organisations), trauma and victimisation). Nevertheless, respondents made clear that CMHTs were only one component to provision of support. Charities, local authorities, community organisations and other NHS services also were reported as playing a role in each domain. Community organisations seemed to be heavily relied upon to meet the social and economic needs of service users across domains. Community organisations or charities were the most cited source of support (compared to the different CMHT roles, other NHS teams, or Local Authorities) in the domains of education and training, community support, social security, income (equal to CMHT care coordinators) and debt. Local authorities were cited as the most frequent source of support for the housing domain (see SI for further results per domain).

## 5. Discussion

Our service evaluation reviewed how social and economic needs are assessed and addressed in mental health services across the UK’s largest mental health provider, South London and Maudsley NHS Foundation Trust (SLaM). Our findings highlight the difficult picture faced by people in contact with mental health services, as well as staff working in this setting. Services are underfunded, staff workloads are high, and responses are oriented towards crises rather than prevention. As such, meeting the social and economic needs of people in contact with services are often deprioritised in service design, making it difficult for staff to respond effectively to the needs they observe. Nevertheless, our case studies illustrate excellent models of how services may better address social and economic needs.

The degree to which social and economic needs are assessed by services varied across different social and economic domains. In each of the ten domains we asked about, over 60% (n = 16) of respondents indicated that needs were always or often assessed. Needs relating to social aspects of life were the most consistently assessed, including family relationships, social connectedness, and community support. Needs relating to education, income and trauma were the least consistently assessed. The only domains that received any reports of a complete lack of assessment were education and employment, although most respondents did report that their teams assessed needs in these domains. This data indicates that CMHTs should have a reasonable understanding of the challenges faced by those in contact with them although improvements are required to increase consistency of assessments of social and economic needs.

Respondents highlighted the lack of adequate support available across all ten social and economic domains, highlighting that the needs of people with mental health problems are currently not being met by services. This emphasises that social and economic needs are deprioritised by health services, despite social interventions being recommended by national guidelines (e.g. IPS, NICE) as a core and necessary component to responding to mental ill-health. Quality of support available was also highlighted as a concern. For example, with respect to employment it is access to meaningful employment which is important for social inclusion, rather than to jobs which may not be suitable and may further negatively impact mental health. Another crucial issue raised was the lack of support available for trauma – this is particularly concerning with respect to care for people with severe mental illness (SMI) such as psychosis, as trauma is highly prevalent in this group (Hardy et al., 2024; Sweeney et al., 2016). Respondents highlighted many unmet needs beyond those related to the ten described domains, for example, support with activities of daily living, sex and relationships, and healthy eating.

In all domains, service users were reported to be supported by a patchwork of teams and groups. This opens up the possibility of the ‘postcode lottery’ as, for example, some geographical areas have fewer third sector organisations than others, and local authorities will be less well-resourced in some areas compared with others. Indeed, some of the services described in our case studies were only available to individuals living within certain boroughs. Further, the geographical areas with the strongest patchwork of support might not correspond to where the level of need is highest – this will require further investigation. The idiosyncratic patchwork of needs, services and third sector organisations in different boroughs may mean that projects which are successful in one area may be hard to replicate in other boroughs with unique patterns of needs and support.

Respondents listed many services and forms of support beyond statutory mental health services and local social services. In many domains, VCSEs were cited as the most common source of support. This reliance on VSCE sector reflects that meeting social and economic needs are deprioritised within services – and perhaps not seen as essential components of health – despite recommendations showing that social interventions should be provided through NHS-commissioned services (e.g. IPS, NICE). VCSEs provide essential support to many people with SMI who are not supported by CMHTs, and many service users trust these organisations to a far greater extent than statutory services, due to long histories of good relational care by VCSEs on the one hand and harmful experiences with statutory services on the other (Mental Health Social Care Policy & and Oversight Group, 2022). Therefore, this patchwork of support with a range of support provided outside the NHS or Local Authorities is crucial for many service users. This raises further concerns as VCSE organisations are often precariously funded and many core social inclusion services are therefore vulnerable to being undermined. Further, these organisations play a particularly important role for people with mental ill-health from further marginalised groups – for example, the Patient and Carer Race Equality Framework (PCREF) emphasises the importance of mental health trusts working with local organisations to meet needs (NHS England, 2025).

The sheer range (if not depth) of types of support and services available highlights the complexity of the landscape of social intervention for people with mental health problems. Service users are unlikely to be aware of the range of support which might be available, and, given that social inclusion is often not the priority of care coordinators, service users may therefore miss out on the support which is available. This suggests the need for specialists to assist service users in navigating this complex system. Some groups are working to provide service users with information about support with social inclusion needs in the catchment area (e.g. Lambeth COIN); however, the challenge of making as many service users as possible aware of such resources remains.

This service survey has several limitations. Firstly, although we recruited participants from all constituent boroughs of SLaM, the small respondent sample size indicates that our results likely do not give a full picture of the landscape of how social needs are assessed and addressed across SLaM. We are unable to determine the representativeness of our sample given the lack of information regarding staffing of the roles of participants we recruited in SLaM. Only a few care coordinators took part in this survey, and we therefore may not have accurately captured their work in this area. Care coordinators typically have very high workloads and therefore participating in research is not a priority: an indicator that there may be limited potential to increase care coordinator’s engagement in social inclusion processes given current capacity levels. Nevertheless, we recruited from a large variety of community teams and involved respondents with many different job roles and titles. Secondly, our convenience sampling method of recruiting participants constitutes a source of bias in our results. We may have recruited staff who are already more passionate about social inclusion and therefore we may have overestimated the degree to which social needs are assessed by teams. Sampling bias may have also resulted in answers in some social domains being more complete than others. We note that our case studies are also convenience-sampled and as such, these examples of good practice are not exhaustive.

Overall, the findings presented here show clear evidence that, despite staff being aware of the high level of needs and attempting to respond, social and economic needs are currently not being adequately met by providers in one of the largest mental health services in the UK. This is likely replicated across services in the UK, as mental health trusts face enormous pressures and social and economic needs are deprioritised. We note that this picture of service provision provides the staff perspective, and it is important to compare these findings to accounts from service users. Nevertheless, these results mirror and support evidence from qualitative studies and testimonials of with people with lived experience of mental health problems.

## Data Availability

All data produced in the present study are available upon reasonable request to the authors

## Author approval

All authors have seen and approved the manuscript.

## Supplementary data

Supplementary information (SI) is available online at medRxiv.

