## Supplementary_Information for "How are social and economic needs assessed and addressed in mental health services? A service evaluation of the largest mental health trust in the UK"

### Aims in full

We aimed to:

iv) Explore the extent to which social and economic needs of people in contact with mental health services are routinely assessed by teams;
iv) Understand what services and support are provided in relation to the social and economic needs of people in contact with mental health services;
iv) Understand the issues and gaps in provision of support with social and economic needs;
iv) Identify whether available support is provided directly by community mental health teams or via referral to others.

In tandem, relevant to aim ii, we aimed to provide in-depth case studies of some examples of excellent practice in this area.

### Methods: Survey full description

The codeveloped service audit survey comprised 41 questions and took approximately 10-15 minutes to complete. The questions were categorised into four parts. The first obtained basic employment information about the participant role (job type, type of team, caseload, caseload guidelines, number of team members). The second asked respondents to indicate how often (always/often/sometimes/never) their team assessed service users’ needs across ten different life domains (employment, education and training; social participation and connectedness; family relationships; community support; social security; debt; income; housing; trauma). The third section comprised a free-text entry form for respondents to list the types of support provided within each of these 10 domains and a multiple choice question concerning who provides this support: “CMHT– generic/ whole-team responsibility/ CMHT – specialist/ CMHT – care coordinator/ CMHT – peer worker/ CMHT – other/ Other NHS services/ Charity/ Local authority/ Other community organisations/ Other”. Participants who selected any response with “other” to these questions were asked to specify to whom they referred; they were also given the opportunity to list any domains of social support that were not covered in the domains listed. The final part of the survey asked respondents about what social needs of people with mental health problems are currently not being met by services; and what evidence they use to inform how they deliver support.

### Case studies

#### Lambeth Vocational Services

**About Lambeth Vocational Services**

Lambeth Vocational Services supports social inclusion for people with mental health problems across the domains of education, employment, training, volunteering. They provide person-centred support to enable individuals to find their own way forward and engage in vocational activities that are important to them. The service also helps service users navigate the welfare benefits and housing systems, as the majority face problems in these domains, which need to be addressed before progress can be made with supporting vocational activities. The service is open to working-age adults who are in contact with mental health services and is accessible through self-referral or referral by a health professional.

A number of projects are facilitated by Lambeth Vocational Services, including community Occupational Therapy; Vocation Matters, a user-led service providing one-to-one support to help people achieve their vocational goals; Community Opportunities Information Network, a resource providing information about local employment, education, leisure, arts, sports and volunteering; and Clean and Care, an employment support project providing paid employment training for people using Lambeth Vocational Services.

The team at Lambeth Vocational Services have published many papers documenting their development as well as researching outcomes achieved for service users. One such qualitative research paper explored the experience of seven service users to examine the features of the service which enable people to achieve their vocational goals. This revealed a range of consistent themes upon which the authors developed a model of validating and invalidating conditions which allow service users to be empowered to find their own way forward. Drawing on this evidence, Lambeth Vocational Services focusses on offering time-unlimited support, building trusting relationships between staff and service users, finding activities that matter to service users, and offering peer support.

**Development and successes**

The founding team began work to establish Lambeth Vocational Services in 2006. They aimed to address the critical lack of social inclusion for people with mental health problems, as emphasised by research finding that 88% of service users were unemployed, yet only 8% received occupational care planning (unemployment: 77% of out-patients, 78% of people on acute wards and 96% under CMHTs; n=267 total, where the most common diagnosis was schizophrenia, n=136) (Bertram & Howard, 2006). Based on consultations with, and facilitated by, service users, the team understood the need for person-centred, co-produced services to promote social inclusion in the context of the high prevalence and severity of adversities service users face, such as poverty and trauma.

The service began with a small occupational therapy pilot team and evolved into a fully-fledged vocational service supporting a significant caseload achieving good social inclusion outcomes for many. The team has successfully secured income for the service in many ways, including through engaging supportive commissioners, obtaining large commercial contracts for the employment creation component of the service, and a partnership with the estates department within the NHS. This success has been achieved despite the many challenges faced over the years, not least the significant budget cuts due to austerity measures, which have limited the reach of their work.

#### Southwark Community Welfare Team

**About SCWT**

The Southwark Community Welfare Team (SCWT) stands as a unique non-statutory service within the Trust, offering expert welfare benefits advice, casework and advocacy to approximately 15 community mental health teams in the Southwark Directorate. The small but dedicated team has expertise in various domains including healthcare and law.

**Key aims**

Primarily, the team aims to improve the financial situation and housing security of service users living within Southwark, thereby tackling the social determinants of mental health problems. Their specialist advice helps service users to navigate often opaque and complex welfare systems facilitating much-needed financial safety and security. The team works to ensure that service users receive the social security they are entitled to, and they are often able to bring service users out of poverty by doing so. Once this basic foundation is in place, service users are better positioned to begin to address other crucial social needs, including meaningful vocation, and participation in social activities. As such, the expert advice offered at SCWT enables service users to actively engage with their community, gain independence and live an overall healthier lifestyle, thus aiding their psychological recovery and equipping service users to remain well, live in the community, prepare for employment, and reduce hospital admissions long-term.

**Successes**

The SCWT service has become even more crucial in response to the ongoing cost-of-living crisis which has had widespread social and economic effects on service users within the Trust catchment. To demonstrate the scale of need, in the first three quarters of 2024 alone, SCWT received around 300 referrals from community mental health services. This need is expected to increase greatly over the coming months as changes in the social security system, comprising the move to Universal Credit, will affect most service users who receive benefits. This highly vulnerable group will often require significant support to navigate this change, which will often involve undergoing further benefit assessments. There is extensive evidence documenting the stress such assessments have on the wellbeing of people with mental health problems and the team at SCWT work hard to minimise potential harm to ensure service users receive the financial support they need.

Since the service was established, SCWT have also built effective partnerships with various secondary care and third-sector services to offer more integrated support such as Southwark Council’s adult social care teams, financial inclusion team and housing benefit assessors, as well as Southwark Jobcentres, debt charities and supported accommodation providers. These partnerships facilitate effective joint workings and, if appropriate, seamless signposting to ensure a smoother journey for service users seeking specialist welfare advice and casework.

By working together with both healthcare professionals internally and a range of relevant third-sector organisations, SCWT has built excellent partnerships to provide an integrated care service at SLaM while embodying Trust values by promoting excellence in mental healthcare. The team at SCWT work to improve the care provided by the Trust by contributing to a more holistic package of care, where social needs – rather than solely medical or psychological needs – are addressed by trained professionals. To further spread awareness of the important work done within SCWT, there are regular sessions available for clinicians within SLaM to learn more about the functions of the SCWT alongside a simple system for staff to make referrals for service users who might need assistance with debt or welfare advice. Due to their well-established links with SLaM clinicians, they are also able to work with service users who are harder to engage with making theirs a unique service which has no equivalent in the community.

#### Psychological Interventions Clinic for Outpatients with Psychosis (PICuP)

**About PICuP**

PICuP is a psychology-led service based at the Maudsley Hospital, specialising in psychological therapies for people with psychosis. They offer psychological interventions for people with distressing positive symptoms of psychosis, including people with bipolar disorder and PTSD diagnoses, as well as those with a history of psychosis whose main difficulties are secondary emotional problems.

Alongside this work, PICuP team members are working to improve social circumstances and reduce social isolation of people with psychosis and their carers through Peer Support Service and Family and Friends Support Service.

**Peer Support Service**

The Peer Support Service is led by a Peer-Recovery lead, who was an ex-client of the PICuP service. It has existed for around ten years and currently comprises 2-5 trained volunteers, supervised by the Peer Recovery lead, who all have recent experience of receiving therapy at PICuP.

The peer support workers at PICuP improve social inclusion of clients in a number of ways. Firstly, they offer one-to-one support for any person with lived experience who attends PICuP. This may include providing emotional and social support to PICuP clients between therapy sessions. Through sharing their own experience of therapy, peer supporters help clients to become familiar with the process while they are on the waiting list, reducing anxiety and avoidance in relation to the therapy process. They also support them to access and engage with therapy through offering practical assistance, for example by accompanying clients to appointments at the clinic when in-person attendance is an obstacle to accessing the help they need.

In addition to offering individual support, the peer support team at PICuP run a weekly group meet-up, alternating between a social group and an arts group. The social group comprises a range of events, from meeting for coffee to visiting museums and galleries together. Importantly, this focusses on free events to ensure the social group is as accessible as possible to all service users.

Peer support at PICuP is available to clients prior to starting and after they have been discharged from therapy. This continuity of support is vital and relatively rare in the mental health care system, where many service users often find themselves without any support after therapy is completed. Participants who continue to engage are never discharged from the peer support service and can continue to be members of the group for as long as they like.

The peer support and involvement initiative has won multiple awards, including The Psychology and Psychotherapy Service User Involvement Award 2015, the Quality, Excellence & Learning Award 2017, The Lammy Awards 2018, SLaM Transforming Lives Award 2018, and The Primary & Community Mental Health Care Award 2018. A research project is underway to determine the representation of marginalised groups within the attendees of the peer support service, as well as to explore qualitatively the experiences they have of the service. Anecdotally, PICuP staff have highlighted that the peer support service is seen as an invaluable source of support for many people and that the Peer Support Lead is highly skilled at providing life-changing and life-saving support to so many PICuP clients. Correspondingly, demand for peer support at PICuP is very high.

Despite a great deal of success, the peer support service faces a number of challenges, particularly with regards to accessing sufficient resources to meet demand. Increased resources might allow for trained volunteers to be paid for the essential work they do, as well as increase the capacity of the team to provide this support to more service users. For example, the art group is hugely popular with service users however capacity is limited due to funding for materials as well as demand on staff time.

**Family and Friends Support Service**

During the COVID-19 pandemic, staff at PICuP identified an urgent need to increase support for the family, friends and carers of clients attending the clinic. The pandemic heightened demands on the family and friends of people with psychosis at a time where formal and informal social support was harder to access. The recognition of the vital support that family and friends play in maintaining optimal support for clients led to the inception of the Family and Friend Support Service.

The service provides 6 weekly sessions to loved ones of PICuP clients. These confidential 50-minute sessions take place either in-person, over the phone, or online, and with consent from the client are offered to each identified carer within the service. The support provided draws on principles established from family intervention research, and employs materials co-produced with carers. The sessions offer the space for carers to share their experiences, learn more about psychosis and bipolar disorder, receive support with relapse prevention and crisis planning, while also working towards individualised goals to improve their own wellbeing. Staff also signpost attendees to sources of additional support where needed. Routine Outcomes Measures (ROMs) are administered pre and post to ensure objective evaluation of the service.

The family and friends service has become a vital part of PICuP, receiving excellent feedback from recipients of this support. It won the 2024 Mental Health Award in the ‘support during pandemic’ award. This year the team were awarded a grant from Change Makers at the Maudsley Charity to further develop the service to include peer support for carers. This peer support project is being coproduced with carers who have attended the Family and Friends Support Service themselves and is due to extend the vital support family, friends and carers receive from the service.

#### Culturally Appropriate Peer Support and Advocacy Service

**About CAPSA**

CAPSA is delivered by Black Thrive CIC ‘Lambeth’ and is funded by Lambeth Living Well Network Alliance. CAPSA provides peer support and advocacy to Black communities in Lambeth who access secondary mental health support within Lambeth. This service was established in July 2021 and works across both the community and in-patient services. The service was co-designed by Black Thrive and Black members of the SLaM Patient and Public Involvement Group and is delivered by peer support workers and advocates. CAPSA champions lived experience to ensure that racially minoritised service users are receiving person centred care that focuses on their cultural needs. Due to this, CAPSA is an operational example of the PCREF (**P**atient, **C**arer **R**ace **E**quity **F**ramework -developed by Black Thrive Director) in action.

**Key aims and ways of working**

CAPSA works hard to help service users and their families to feel empowered in their recovery journeys. The team finds new, innovative ways of working to:

- Address the failures of current mainstream services when it comes to providing care for Black communities
- Tackle racial inequalities in mental health services

The CAPSA team is comprised of 4 individuals that provide peer support and advocacy to support a well-rounded service for their clients. All advocates undertake advocacy training to use the statutory frameworks to support clients. Referrals to CAPSA come through clinicians. Once service-users are referred and are taken on by the service as clients, CAPSA works to empower these clients to both self-advocate and seek support in their recovery journey. CAPSA peer advocates work to challenge diagnosis, medication, restrictive practices, and treatments that disproportionately impact Black service users, such as Community Treatment Orders and Seclusion. All CAPSA workers also offer 1-2-1 support in both the community and in-service, which includes liaising with different stakeholders in care, family members, friends and clinicians. Where CAPSA cannot offer official advocacy, such as when clients enter the social care or criminal justice systems, the team can provide emotional support.

**Impact**

CAPSA, a relatively new service, has already gained recognition for its innovative approach to care. CAPSA recently received the HSJ's 2022 'Best Not-for-Profit Working in Partnership with the NHS' award. CAPSA has also collaborated with the Lambeth Hospital Exercise Team and the Professional Football Association to hold a wellbeing event that included a 5 a side football match between each ward and CAPSA team members, followed by a conference attended by the chair of SLaM. During a recent service review, prior CAPSA clients indicated a sense of belonging as well as accountability for both them and the services they use as a result of the involvement of CAPSA members.

### Survey results: Participant roles

Advanced Nurse Practitioner (n=1), Business Manager (n=1), Care coordinators (n= 3), Clinical Psychologist (n=2), Community Practitioner (n=1), Consultant Psychiatrists (n=3), Clinical Site Manager (n=1), General Manager (n=1), Heads of services  (n=2), Liaison Psychiatrist (n=1), NHSP MH Practitioner (n=1), Nursing Associate (n=1), Occupational Therapist (n=3), Senior Psychological Therapist (n=2), Service Manager (n=1), Social Worker (n=1), Support Worker (n=2), Team Manager (n=1).

### Survey results: Teams and caseloads

The median caseload reported by people working in community teams was 25. Guidelines for caseloads varied, for example from 3 cases per day for a clinical psychologist, 5 per day for an Advanced Nurse Practitioner, 12 per day for a consultant psychiatrist; the largest caseload guidelines reported were 25 for a Social Worker and 30 for CMHT staff but the timespan for this caseload was not reported. A significant number of respondents reported that there were no guidelines or that they were not sure or not aware of any guidelines in place (n=14).

Respondents worked in multidisciplinary teams, ranging from 8-31 team members, although 8 respondents did not provide team size. Listed staff roles varied, for example including assistant psychologists, care coordinators and employment support workers.

Coworkers roles in teams reported included: activity coordinators, administration support workers, advance nurse practitioners, assistant psychologists, care coordinators, clinical psychologists, community psychiatric nurses, consultants, doctors, dual diagnosis workers, employment support workers, GP trainees, health and wellbeing trainees, healthcare assistants, Individual Placement and Support workers, locum senior house officers, mental health nurses, metal health care support workers, nurses, occupational therapists, peer support workers, pharmacists, physiotherapists, psychiatrists, reablement support workers, senior therapists, social workers, speech and language therapists, students, support workers, Support, Time and Recovery (STAR) workers, team administrators, team managers, trainee psychiatrists, trainee psychologists, vocational workers, and wellbeing practitioners.

### Survey results: Who provides support? Figures

Employment

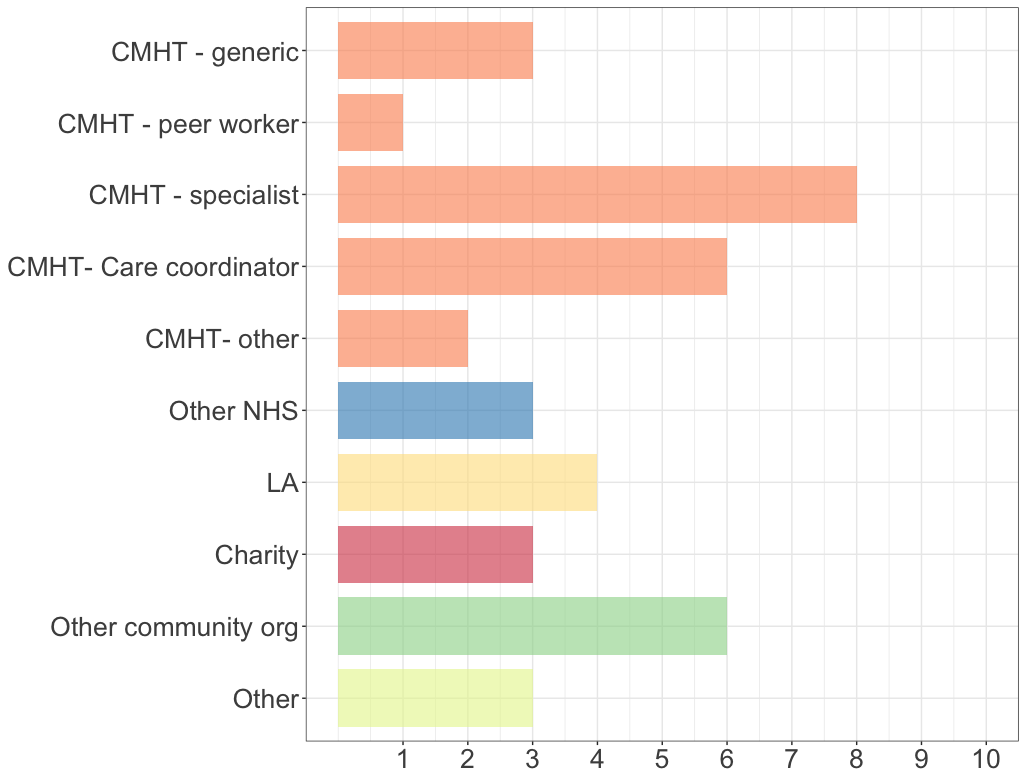

Education and training

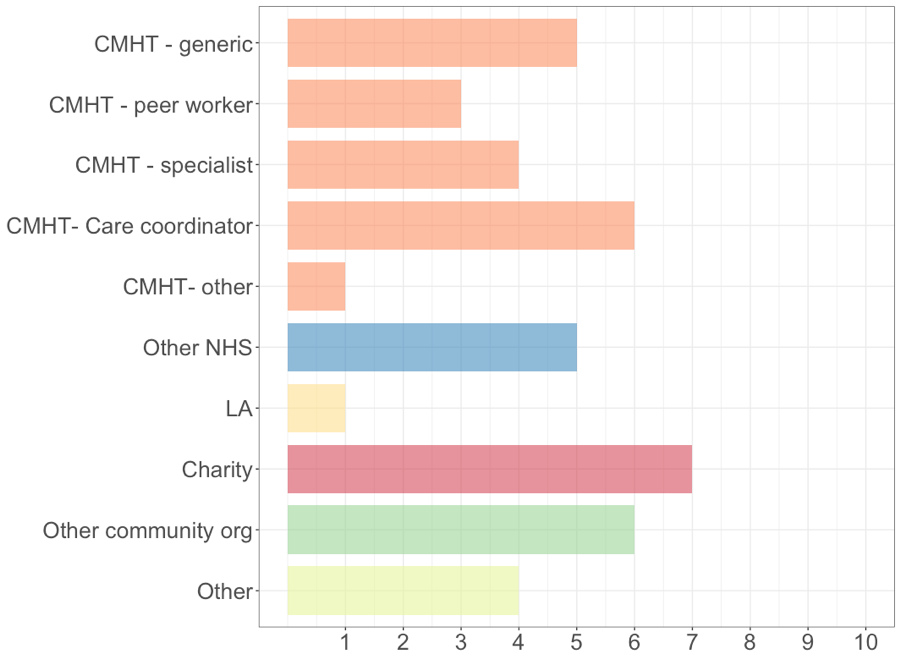

Social participation and connectedness

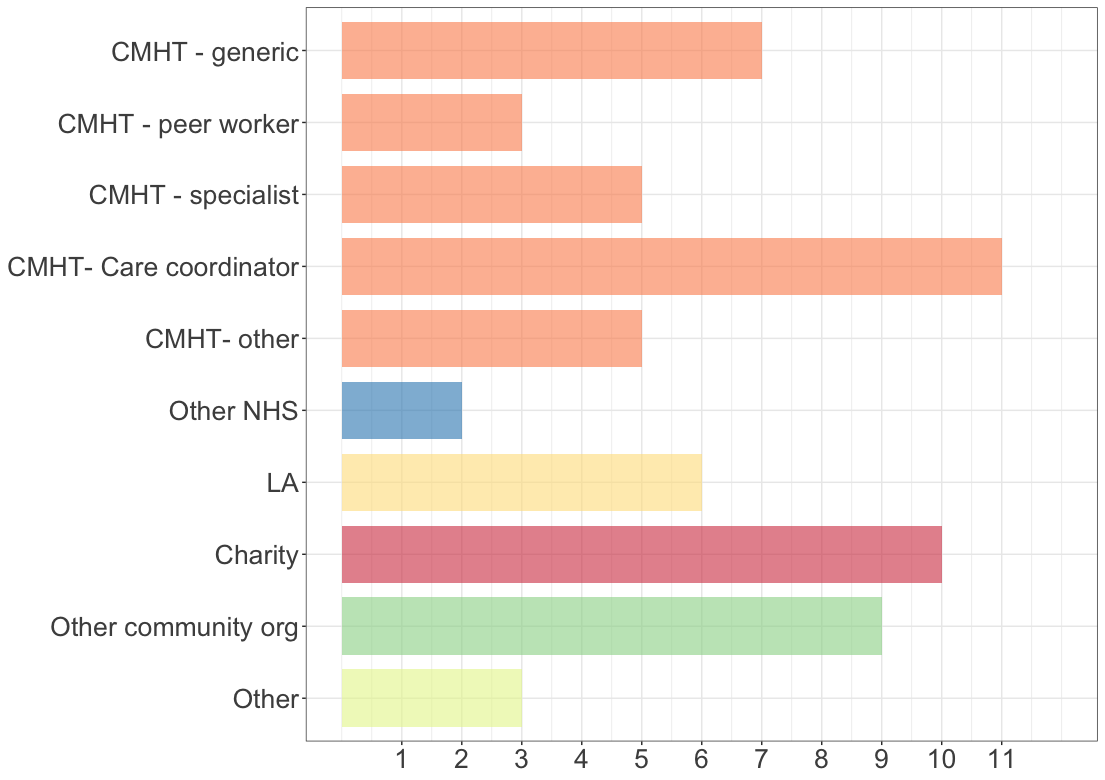

Family relationships

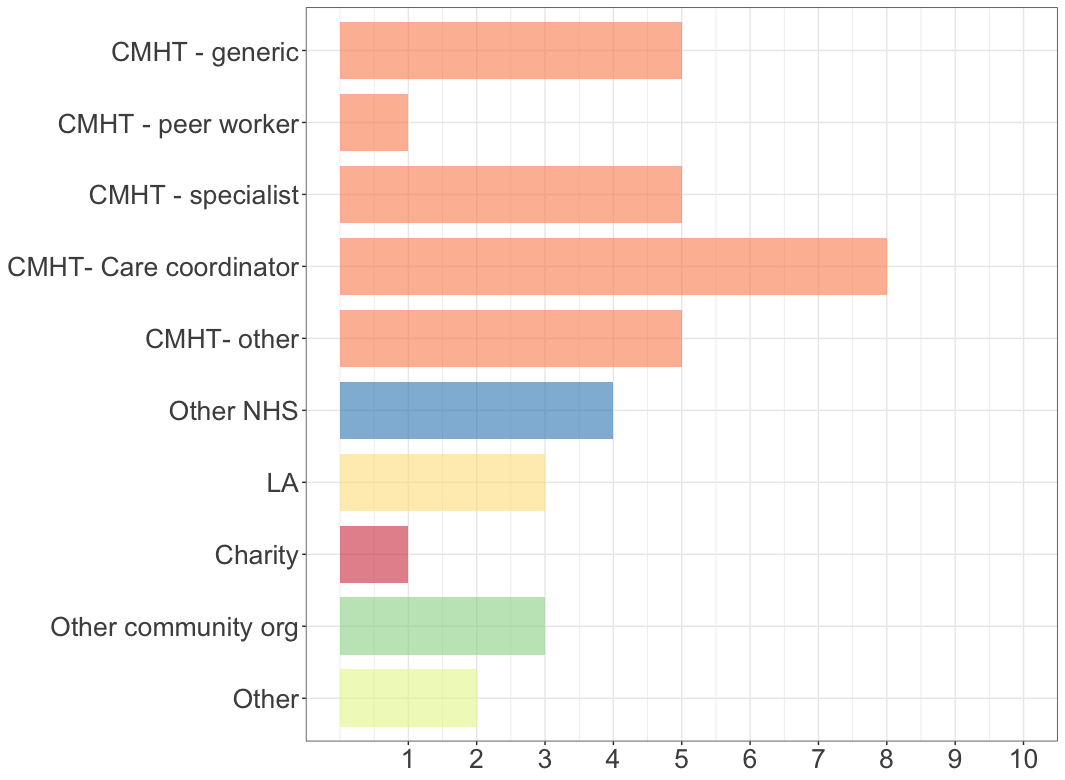

Community support

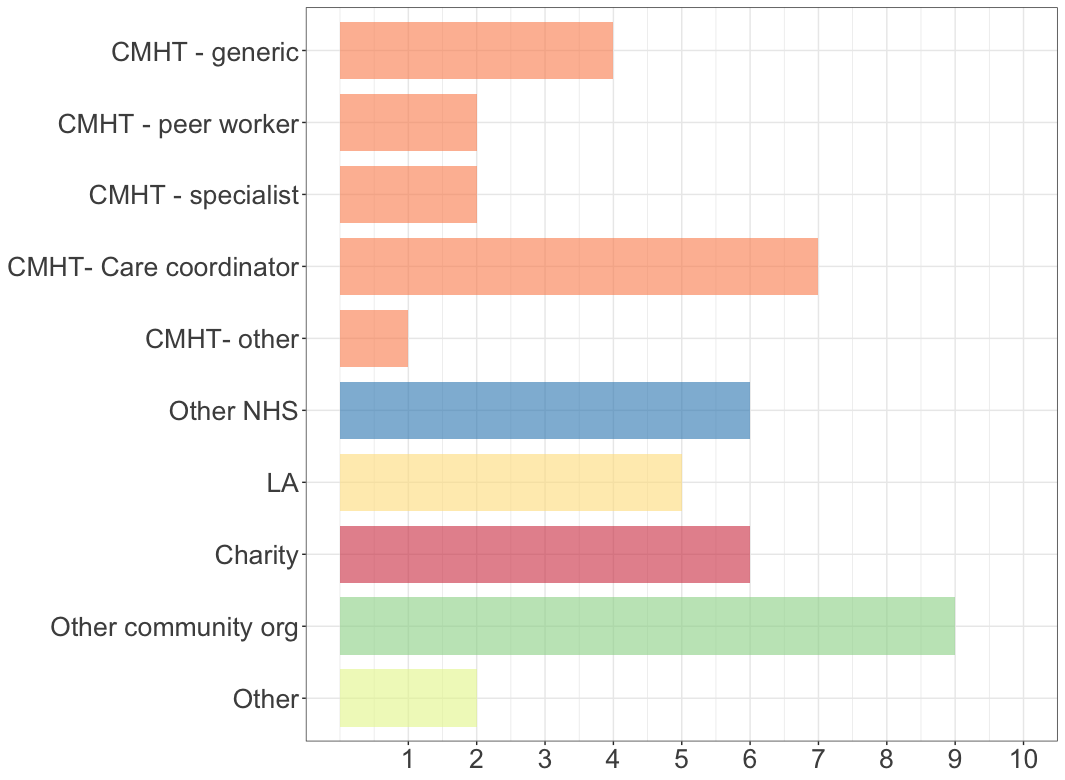

Social security

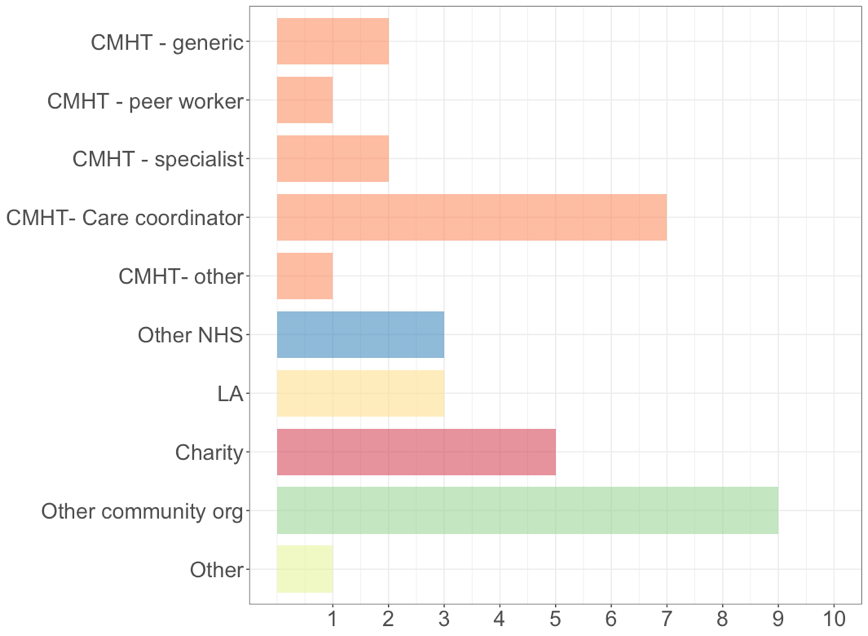

Debt

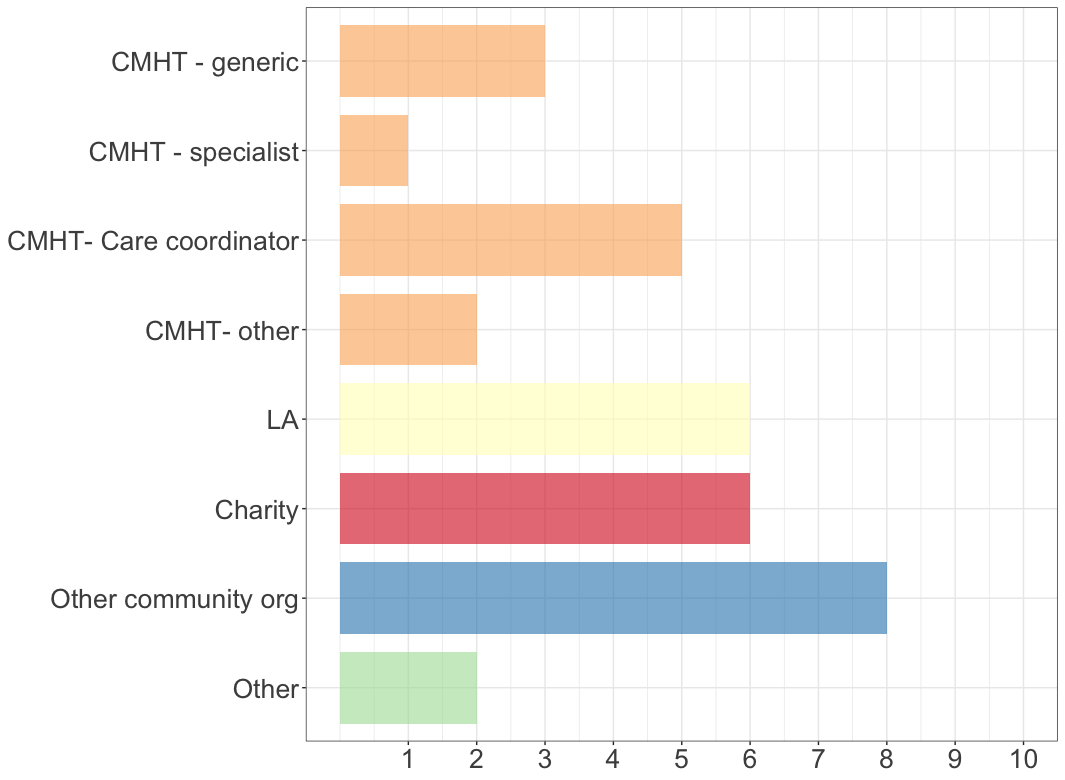

Income

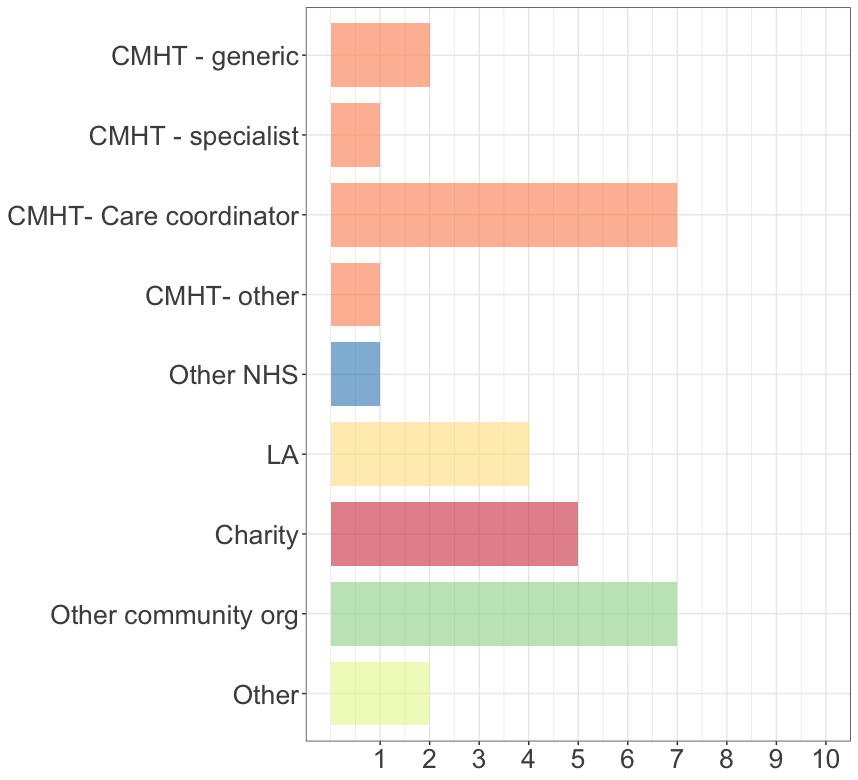

Housing

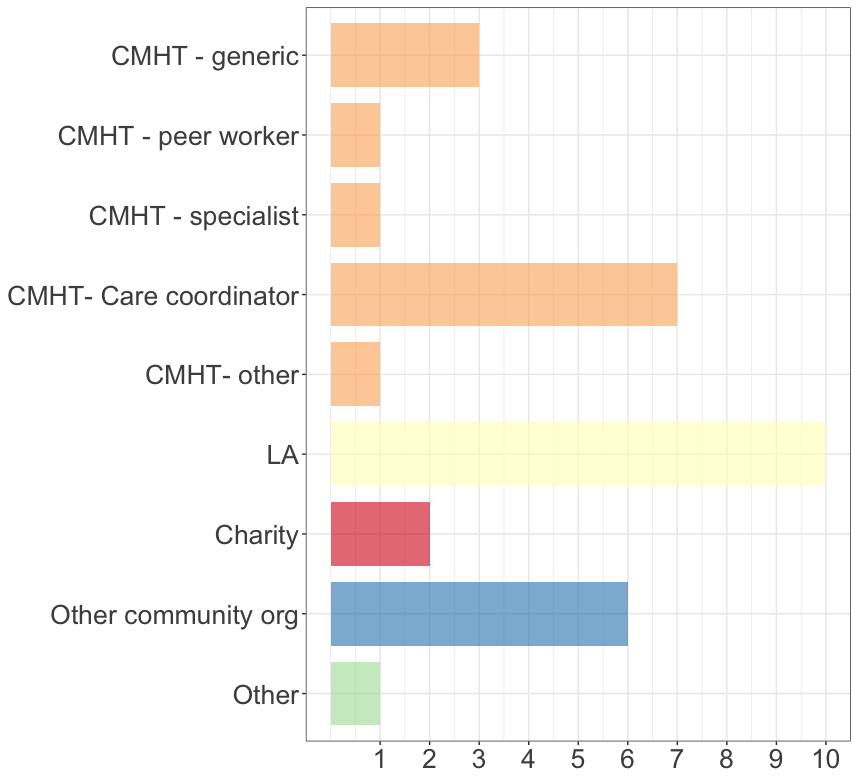

Trauma and victimisation

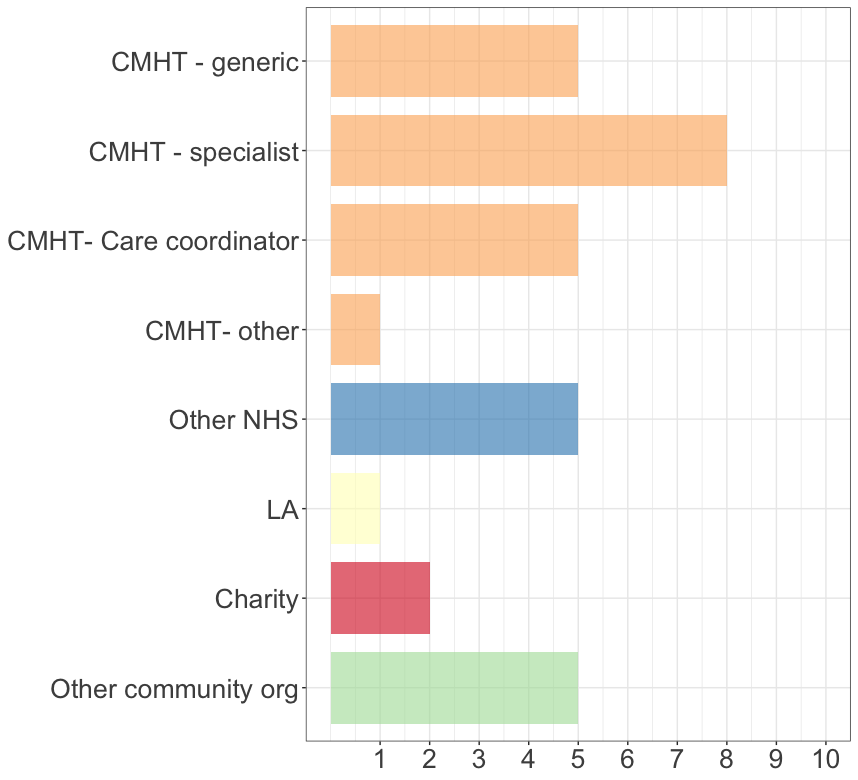
